# Short-chain fatty acid concentrations in the incidence and risk-stratification of colorectal cancer: a systematic review and meta-analysis

**DOI:** 10.1101/2022.03.13.22272319

**Authors:** Ehsan Alvandi, Wilson K M Wong, Mugdha V Joglekar, Kevin J Spring, Anandwardhan A Hardikar

## Abstract

The beneficial role of gut microbiota and bacterial metabolites, including short-chain fatty acids (SCFAs), is well recognized; although the available literature around their role in colorectal cancer (CRC) has been inconsistent.

We performed a systematic review and meta-analysis to examine associations of fecal SCFA concentrations to the incidence and risk of CRC.

Data extraction through Medline, Embase, and Web of Science was carried out from database conception to May 21, 2021. Predefined criteria included human clinical observational studies, while excluding cell/animal model studies, conference proceedings, and reviews. Quality assessment of selected 16 case-control and six cross-sectional studies is reported using PRISMA 2020 guidelines. Studies were categorized for CRC risk or incidence, and RevMan 5.4 was used to perform the meta-analyses. Standardized mean differences (SMD) with 95% confidence intervals (CI) were calculated using a random-effects model.

Combined analysis of acetic-, propionic-, and butyric-acid revealed significantly lower concentrations of these SCFAs in individuals with high-risk of CRC (SMD = 2.02, 95% CI 0.31 to 3.74, P = 0.02). Further, CRC incidence increased in individuals with lower levels of SCFAs (SMD = 0.45, 95% CI 0.19 to 0.72, P = 0.0009), compared to healthy individuals.

Overall, lower fecal concentrations of the three major SCFAs is associated with higher risk and incidence of CRC.

## Introduction

According to the Global Cancer Incidence (GLOBOCAN) 2020 report, colorectal cancer (CRC) is the third-most commonly diagnosed cancer (10% of all diagnosed cancers) and the second (9.4%) leading cause of cancer-related death (Sung *et al*. 2021). It has been estimated that the overall risk of CRC in all age groups will increase 60% worldwide by 2030, leading to more than 1.1 million deaths and 2.2 million new cases (Arnold *et al*. 2017). Colorectal cancer mostly develops from precursor lesions arising from the aberrant proliferation of colonic epithelial cells (colonocytes) and are commonly referred to as adenomatous polyps or colorectal adenomas (CRA) (Dekker *et al*. 2019; L. H. Nguyen, Goel and Chung 2020). It is a heterogeneous disease and environmental factors have a potential impact on the development of CRC, among which diet is a risk factor (Dekker *et al*. 2019; Mármol *et al*. 2017; Wong and Yu 2019). According to several meta-analyses, high consumption of processed and unprocessed meat is related to high CRC risk (Chan *et al*. 2011; Zhao *et al*. 2017), and high fibre intake is suggested as a protective factor against CRC progression and incidence (Gianfredi *et al*. 2018; Nucci *et al*. 2021; Oh *et al*. 2019).

The effect of diet on colonic health is partly mediated through alteration of gut microbiota composition, diversity, and metabolism (S. J. O’Keefe 2016; Wong and Yu 2019). Gut microbiota constitutes the largest community of commensal microorganisms in the body, which mainly resides in the lower small intestine and colon (S. J. O’Keefe 2016; Wong and Yu 2019; Yang and Yu 2018). The gut microbiota-derived metabolites are in constant crosstalk with colonocytes, and short-chain fatty acids (SCFAs) make up a large group of these metabolites (S. J. O’Keefe 2016; Wong and Yu 2019; Yang and Yu 2018).

Short-chain fatty acids are small molecules generated via the fermentation of dietary fibres by gut microbiota. Acetic, propionic and butyric acid constitutes the majority of colonic SCFA content (Alexander *et al*. 2019; Parada Venegas *et al*. 2019) and the beneficial anti-inflammatory and anti-carcinogenic effects of dietary fibres on colonocytes are mediated through these SCFA molecules (Liu *et al*. 2021; van der Beek *et al*. 2017). Among the three major SCFA molecules, butyric acid is also considered as one of the main energy sources for colonocytes (Alexander *et al*. 2019; S. J. O’Keefe 2016; van der Beek *et al*. 2017). Therefore, alteration in SCFA levels could impact the colonic health and predisposition of colonocytes to aberrant proliferation and tumor formation (Liu *et al*. 2021; Parada Venegas *et al*. 2019).

Several studies have assessed fecal SCFA concentration in patients with colorectal carcinoma or adenoma (Boutron-Ruault *et al*. 2005; Bridges *et al*. 2018; C. Y. Chen *et al*. 2021; H. M. Chen *et al*. 2013; Kashtan *et al*. 1992; Lin *et al*. 2016; Lin *et al*. 2019; Monleon *et al*. 2009; Niccolai *et al*. 2019; Ohigashi *et al*. 2013; E. M. Song *et al*. 2018; Sze *et al*. 2019; Torii, Kanemitsu and Hagiwara 2019; Weaver *et al*. 1988; Weir *et al*. 2013; Yusuf, Adewiah and Fatchiyah 2018). However, due to variable results, the conclusive evaluation of SCFA profiles from CRC patients versus healthy subjects is lacking. In addition, other studies have compared SCFA concentration within healthy individuals from various countries and ethnic groups with the highest and lowest prevalence of CRC; although with inconsistent results (Hester *et al*. 2015; Katsidzira *et al*. 2019; S. J. D. O’Keefe *et al*. 2009; Ocvirk *et al*. 2020; Ou *et al*. 2013; Ou *et al*. 2012).

Therefore, systematic analyses designed to better understand the link between SCFA concentration in CRC risk and incidence is highly desired. We aimed to systematically analyse the results of all primary observational human studies, which have measured fecal SCFA levels in at-risk individuals or CRC patients. We divided our analyses on the available evidence into two categories based on (1) CRC risk and (2) incidence. In the CRC risk category, the focus was on at-risk individuals, therefore we included two groups of studies where fecal SCFA concentration is compared between at-risk individuals with colorectal adenoma versus healthy individuals (1a), and individuals at high-versus low-risk of CRC based on the prevalence of the disease between various countries and/or ethnic groups (1b). In the CRC incidence category the focus was on studies that compared the fecal SCFA levels in individuals with CRC versus healthy individuals. Our results underline the potential association of the three major SCFA molecules (acetic, propionic, and butyric acid) with CRC risk and incidence.

## Methods

We used Preferred Reporting Items for Systematic Reviews and Meta-Analyses (PRISMA) 2020 guideline (Muka *et al*. 2020; Page *et al*. 2021) to systematically search and extract data from primary human studies with SCFA measurement in CRC risk or incidence.

### Database search

The Medline, Embase, and Web of Science database search was performed for articles involving human subjects that are in English from database conception until 21^st^ May 2021. The details of the search keywords and strategies utilized in Ovid and Web of Science are available in the **Supplementary Methods** section.

### Eligibility criteria

All the records, including abstracts, were imported to EndNote X9 (Clarivate Analytics, Toronto, Canada). Duplicate records were first removed. The records were then filtered using EndNote’s built-in search tool for the following criteria: (i) searching for concentration*, level*, quanti*, measure*, assess*, evaluat*, estimat*, calculat*, mmol, and µmol as the inclusion criteria to capture studies which reported the SCFA measurement based on these terms. The asterisk symbol (*) applied was to include all the variations of the search terms, and (ii) searching for mouse, mice, murine, rats, conference, ethyl acetate (EtOAc), and phorbol as the exclusion criteria to exclude rodent studies, conference proceedings, and studies that have stated the use of any unrelated chemicals (such as EtOAc and 12-O-Tetradecanoylphorbol-13-acetate). The abstracts of the remaining records were then screened to exclude reviews, methodology, human studies not related to SCFAs in CRC or CRA, non-human studies (i.e., *in vitro* or other non-rodent animal studies), to identify the human studies on SCFA measurement in CRC or CRA. The full text of the remaining (n = 53) records were then screened to include only the observational studies which have measured fecal SCFA concentration. A final set of 22 observational studies qualified for further data extraction and quality assessment for meta-analysis.

### Data extraction and quality assessments

The data and additional details available for analysis (such as study subjects and SCFA levels) from the finalized primary studies were extracted and added to an Excel worksheet. The Newcastle-Ottawa Scale (NOS) (Wells *et al*. 2011) was used as a standard tool for quality assessment of 16 case-control studies in the selection, comparability and exposure categories, to provide a score range between 0-9 (≤ 6, 7-8, and 9 indicate high, medium, and low risk of bias, respectively) (Muka *et al*. 2020). Evaluation of six cross-sectional studies was performed using the Joanna Briggs Institute (JBI) Critical Appraisal Checklist tool (Moola *et al*. 2020), as recommended (L. L. Ma *et al*. 2020).

### Statistical analyses

Review Manager (RevMan) software version 5.4 (Cochrane, Copenhagen, Denmark) was used to analyse the quantitative fecal SCFA concentration data, which were available in 10 of the final 22 observational studies (9 of 16 case-control, plus 1 of 6 cross-sectional studies). The fecal concentration of acetic, propionic, or butyric acid was considered as the subgroups. Before data entry, SEM or 95% CI upper and lower bound values were converted to SD. Due to variation in the reported SCFA concentration units between different papers, standardized mean difference (SMD) was selected as a measure of effect size for each study. The statistical heterogeneity among studies was calculated using Chi^2^ and *I*^2^ tests and a P-value of 0.05 was considered significant (Higgins *et al*. 2021). To normalise the use of different SCFA measurement methods, a random-effects model was applied to analyse the pooled effect size and P-value for each SCFA molecule in each subgroup. One overall effect size and P-value of combined acetic, propionic, and butyric acid were also calculated. In all analyses, the effect size was reported with 95% confidence intervals, and the P-value < 0.05 was considered significant. Furthermore, the fixed-effect model was also applied in the case of non-significant heterogeneity of *I*^2^ < 50 (Higgins *et al*. 2021). All the data conversions, as well as qualitative and quantitative analyses, were validated by the second team member and confirmed by the senior authors.

One study (Ocvirk *et al*. 2020) reported the numeric values of butyric acid concentration and other SCFA molecules in graphs, hence was included in both quantitative (for butyric acid) and qualitative data (for acetic acid, propionic acid and total SCFA). Therefore, 13 of 22 studies in which the fecal SCFA concentration was presented using graphs (with no reported actual values) were considered as qualitative studies (7 of 16 case-control, plus all 6 cross-sectional studies – including *Ocvirk et al. 2020*). The outcome of analyses from these qualitative studies was plotted as stacked bar charts, using Microsoft Excel (ver. 2016; Microsoft Corporation, Redmond, WA, USA).

## Results

### Study selection and quality assessment

The workflow on the identification and stepwise selection of the observational studies is presented in **Figure 1**. Initially, a total of 2000 English language records obtained from searching through the three databases (Medline, Embase, and Web of Science) were imported to EndNote along with their abstracts. After removing duplicate records, the titles and abstracts of the remaining 1373 records were filtered and screened for eligibility as detailed in the Methods section. In total, 1320 records were excluded, of which most were *in vitro* studies. From the remaining 53 human studies, 31 were excluded. Of these 27 were interventional studies, one was an observational study on serum SCFA (Baldi *et al*. 2021), two studies had indistinct grouping (one case-control study with the presence of individuals with adenomatous polyps in the healthy control group (Amiot *et al*. 2015) and one cross-sectional study with no clear definition of CRC high- and low-risk group (Segal *et al*. 1995)), and one retracted observational study (Wang *et al*. 2017).

**Figure 1.**
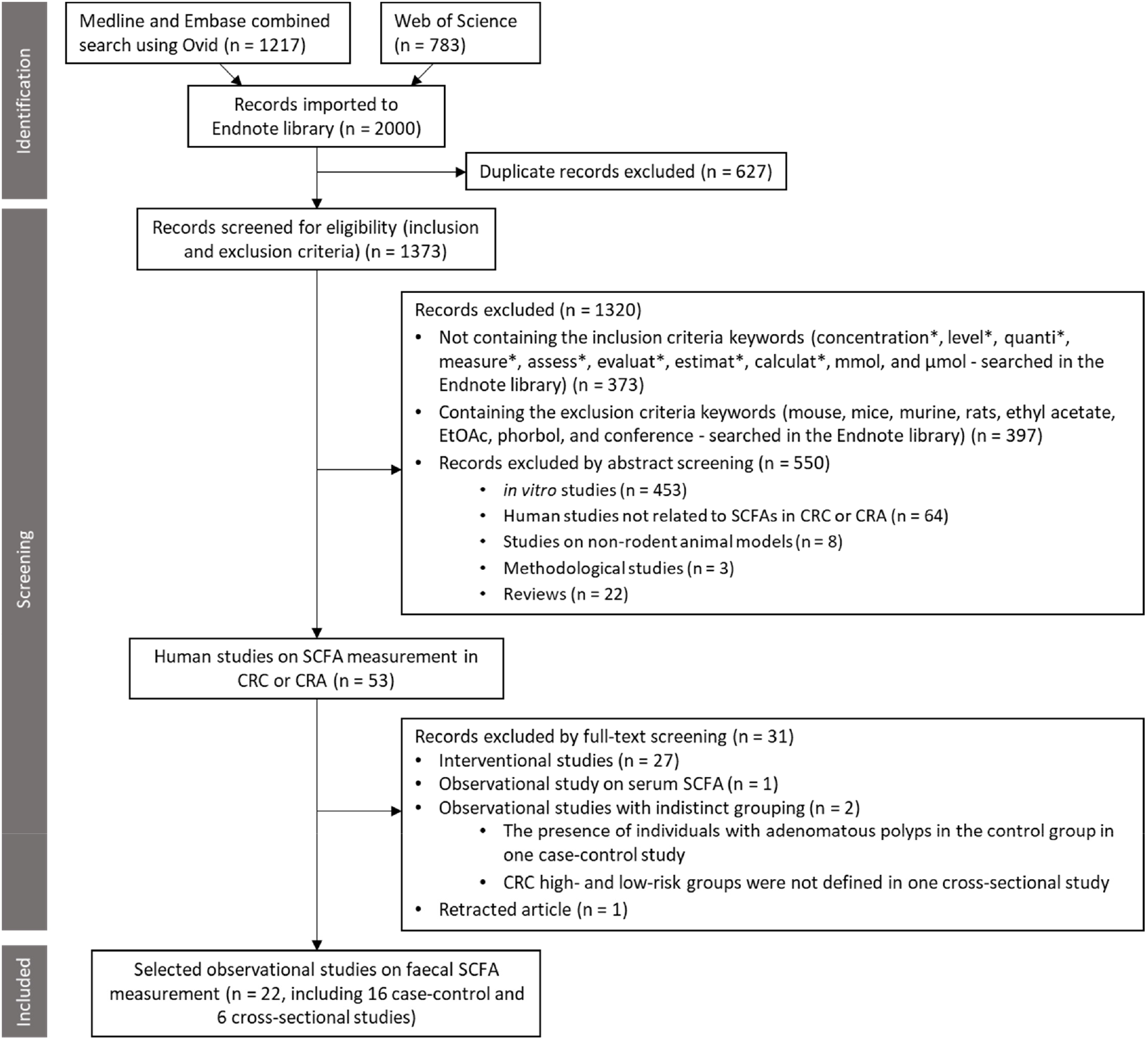
The PRISMA flowchart shows the selection process of the systematic review. The abstracts of all the studies were imported into Endnote from the indicated databases. SCFA: short-chain fatty acid, CRC: colorectal cancer, CRA: colorectal adenoma.

A final of 16 case-control and 6 cross-sectional studies were selected for data extraction and analysis. **Table 1** summarises the characteristics of these observational studies. The results of quality assessment using NOS and JBI tools on case-control and cross-sectional studies are provided in **Supplementary Tables 1** and **2**, respectively.

**Table 1:**
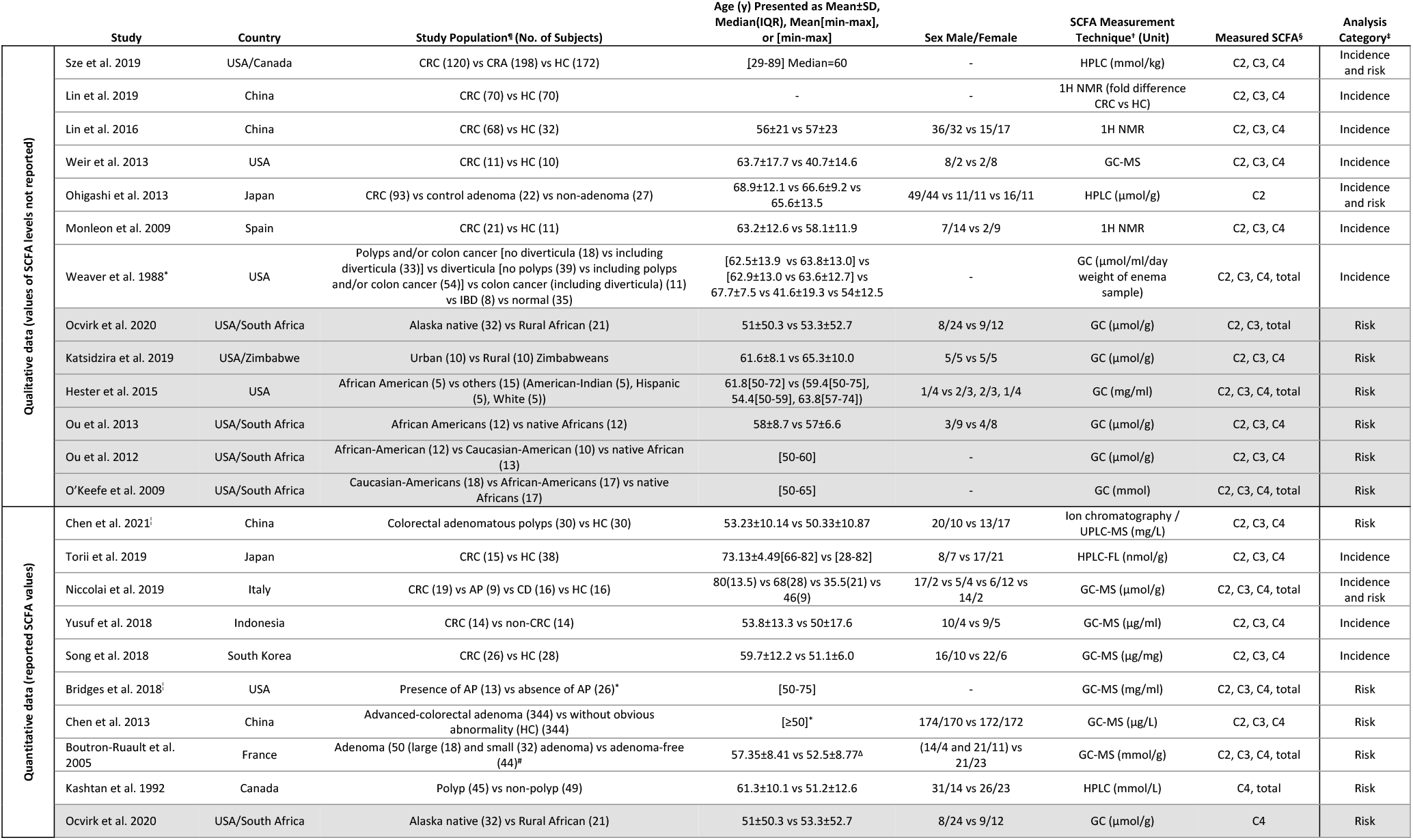
Characteristics of the selected studies. Cross-sectional studies are highlighted in grey, and case-control studies are not highlighted. ^¶^CRC: colorectal cancer, AP: adenomatous polyposis, CD: celiac disease, CRA: colorectal adenoma, HC: healthy controls, IBD: inflammatory bowel disease. ^†^GC-MS: gas chromatography-mass spectrometry; HPLC: high-performance liquid chromatography; FL: fluorescence; 1H NMR: 1H nuclear magnetic resonance spectroscopy; UPLC-MS: ultra-performance liquid chromatography-tandem mass spectrometry; GLC: gas-liquid chromatography. ^§^C2: acetic acid, C3: propionic acid, C4: butyric acid. ^‡^Refer to the text for the definition of CRC risk and incidence category. ^°^Values in this paper were measured on enema samples, not faeces. Therefore, they used in qualitative analysis. ^*^More details are provided in the article. ^¦^Removed from meta-analysis due to insufficient data on SCFA measurement method. ^#^SCFAs were measured in only a subset of these subjects (n = 25 large/small adenoma and n = 23 adenoma-free). ^Δ^Combined values of males and females.

### Stratifications based on CRC risk or incidence

Studies listed in **Table 1** are presented based on the type of data provided (qualitative or quantitative) and CRC risk and/or incidence. Among the 16 case-control studies (not highlighted in **Table 1**), 8 studies comparing CRC cases and healthy control subjects were allocated to the CRC incidence category, 5 studies comparing individuals with CRA and healthy controls assigned to the CRC risk category, and the remaining 3 studies were included in both incidence and risk categories since they compared CRC patients, CRA individuals and healthy subjects. All 6 cross-sectional studies (highlighted grey in **Table 1**) comparing populations with high-versus low-risk of CRC were allocated to the risk category. Therefore, the CRC incidence and risk category included 11 and 14 studies, respectively (**Table 1**). For each study, the details of the measured SCFA and CRC risk and/or incidence grouping are provided in **Supplementary Table 3**. Some studies reported total SCFA concentration in addition to the individual (acetate, propionate, and butyrate) SCFAs.

The primary studies analysed in this systematic review were performed in various countries and ethnic groups. Age was matched in some of the studies (Boutron-Ruault *et al*. 2005; Lin *et al*. 2019; S. J. D. O’Keefe *et al*. 2009; Ou *et al*. 2013; Ou *et al*. 2012; E. M. Song *et al*. 2018), although the male-to-female ratio was not similar between the study groups in most studies (**Table 1**). The SCFA concentrations were measured using different techniques, such as gas chromatography, liquid chromatography, gas-liquid chromatography and ^1^H nuclear magnetic resonance spectroscopy.

### Data analyses

The meta-analysis of the quantitative data extracted from the 10 selected studies (Boutron-Ruault *et al*. 2005; Bridges *et al*. 2018; C. Y. Chen *et al*. 2021; H. M. Chen *et al*. 2013; Kashtan *et al*. 1992; Niccolai *et al*. 2019; Ocvirk *et al*. 2020; E. M. Song *et al*. 2018; Torii, Kanemitsu and Hagiwara 2019; Yusuf, Adewiah and Fatchiyah 2018) are presented in **Figure 2**. In the risk category (**Figure 2A and B**), two studies (Bridges *et al*. 2018; C. Y. Chen *et al*. 2021) were excluded from the meta-analysis due to the lack of sufficient details of the methods used for SCFA measurement from stool samples. In CRC risk meta-analysis, the effect size of each of the three SCFAs was not statistically significant, however, their combined effect size was significantly higher in low-risk compared to high-risk CRC (SMD = 2.02, 95% CI 0.31 to 3.74, P = 0.02, **Figure 2A**). The effect size of total SCFA concentration was not statistically significant in the low- vs high-risk group **(Figure 2B**).

**Figure 2.**
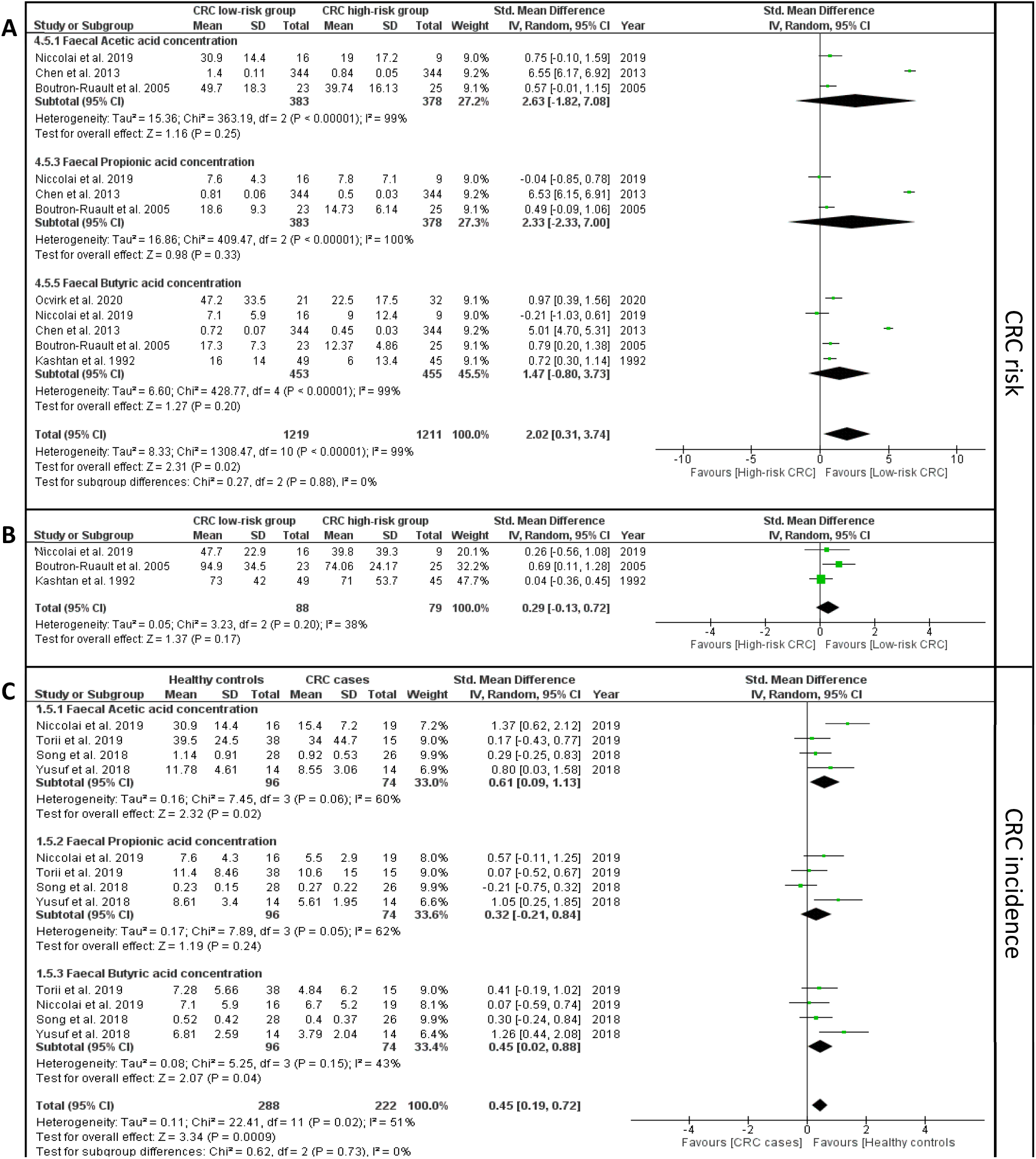
Forest plots representing the meta-analyses of the fecal concentrations of **A)** acetic, propionic, and butyric acid in CRC risk category; **B)** total SCFA in CRC risk category; and **C)** acetic, propionic, and butyric acid in CRC incidence category. Note that in **B**, the total SCFA indicates the collection of all the SCFA molecules - not only acetic, propionic, and butyric acid.

In the CRC incidence analysis (**Figure 2C**), the fecal concentrations of acetic acid (SMD = 0.61, 95% CI 0.09 to 1.13, P = 0.02) and butyric acid (SMD = 0.45, 95% CI 0.02 to 0.88, P = 0.04) were significantly higher in the healthy control compared to CRC cases. In addition, the combined effect size of acetic, propionic, and butyric acid remained significant between CRC cases and healthy controls (SMD = 0.45, 95% CI 0.19 to 0.72, P = 0.0009, **Figure 2C**).

Furthermore, the *I*^2^ heterogeneity index was in the “moderate” range (30% to 60%) (Higgins *et al*. 2021) for the meta-analysis of total SCFA concentration in CRC risk (**Figure 2B**) and butyric acid in CRC incidence (**Figure 2C**) category. Therefore, we performed another meta-analysis using the fixed-effect model on the same data instead of the random-effect model presented in **Table 2**. This resulted in a more pronounced difference in butyric acid concentration between CRC cases and healthy controls (SMD = 0.42, 95% CI 0.1 to 0.74, P = 0.009). The results of the fixed-effect model meta-analyses are presented in **Supplementary Figures 1 and 2**, respectively, and the findings of all quantitative meta-analyses are summarised in **Table 2**.

**Table 2.** Summary of the outcomes of each meta-analysis. Significant P values of the effect size are in bold. ^¶^Combined effect size of acetic, propionic, and butyric acid. Note that the total SCFA indicates the collection of all the SCFA molecules - not only acetic, propionic, and butyric acid.

| | Measured SCFA | Number of Studies | Heterogeneity ($I^2$ %, P value) | Statistical Model | Effect size (SMD [95% CI], P value) |
| --- | --- | --- | --- | --- | --- |
| CRC risk | Acetic acid | 3 | 99, < 0.00001 | Random effect | 2.63 [-1.82 to 7.08], 0.25 |
|  | Propionic acid | 3 | 99, < 0.00001 | Random effect | 2.33 [-2.33 to 7.00], 0.33 |
|  | Butyric acid | 5 | 99, < 0.00001 | Random effect | 1.47 [-0.80 to 3.73], 0.2 |
|  | Combined<sup>†</sup> | 11 | 99, < 0.00001 | Random effect | 2.02 [0.31 to 3.74], **0.02** |
|  | Total SCFA | 3 | 38, 0.2 | Random effect | 0.29 [-0.13 to 0.72], 0.17 |
|  | Total SCFA | 3 | 38, 0.2 | Fixed effect | 0.25 [-0.05 to 0.56], 0.11 |
| CRC incidence | Acetic acid | 4 | 60, 0.06 | Random effect | 0.61 [0.09 to 1.13], **0.02** |
|  | Propionic acid | 4 | 62, 0.05 | Random effect | 0.32 [-0.21 to 0.84], 0.24 |
|  | Butyric acid | 4 | 43, 0.15 | Random effect | 0.45 [0.02 to 0.88], **0.04** |
|  | Combined<sup>†</sup> | 12 | 51, 0.02 | Random effect | 0.45 [0.19 to 0.72], **0.0009** |
|  | Butyric acid | 4 | 43, 0.15 | Fixed effect | 0.42 [0.1 to 0.74], **0.009** |

Qualitative analysis was carried out on the studies which reported lower, higher or no changes to the concentration of SCFAs between high-risk CRC (for risk category) or CRC case (for incidence) and low-risk or control, respectively (Hester *et al*. 2015; Katsidzira *et al*. 2019; Lin *et al*. 2016; Lin *et al*. 2019; Monleon *et al*. 2009; S. J. D. O’Keefe *et al*. 2009; Ocvirk *et al*. 2020; Ohigashi *et al*. 2013; Ou *et al*. 2013; Ou *et al*. 2012; Sze *et al*. 2019; Weaver *et al*. 1988; Weir *et al*. 2013) (**Supplementary Figure 3**). In the risk category, more studies (66.7%) reported significantly lower concentrations of fecal acetic, propionic, and butyric acid as well as total SCFA in individuals at high risk of CRC. In the incidence category, more studies (69%) reported significantly lower concentrations of fecal acetic and butyric acid in CRC patients compared to healthy controls. However, the number of studies reporting no significant difference in the propionic acid was the highest in the incidence category. Overall, our qualitative analysis (**Supplementary Figure 3**) corroborates with the meta-analysis results (**Figure 2**).

## Discussion

For more than three decades, *in vitro*, animal, and human studies have identified numerous potentially beneficial anti-inflammatory and anti-carcinogenic roles of SCFA molecules in gut health and colonic diseases (Alexander *et al*. 2019; Liu *et al*. 2021; Parada Venegas *et al*. 2019;

M. Song, Chan and Sun 2020; van der Beek *et al*. 2017; Wong and Yu 2019). In addition, several meta-analyses (**Supplementary Table 4**) have assessed the role of colonic microbiota (Borges-Canha *et al*. 2015), non-digestible carbohydrates (Rao *et al*. 2021) and dietary fibre in colorectal carcinoma (Gianfredi *et al*. 2018; Y. Ma *et al*. 2018) or adenoma (Nucci *et al*. 2021; Oh *et al*. 2019) as well as the alteration of SCFAs in irritable bowel syndrome (IBS) (Sun *et al*. 2019), or inflammatory bowel disease (IBD) (Zhuang *et al*. 2019).

This systematic review and meta-analysis were conducted on 22 studies to better determine the potential association between fecal SCFA concentration and CRC risk and incidence. The combined mean difference of acetic, propionic, and butyric acid in the CRC risk category analysis revealed a significantly lower concentration of these SCFAs in individuals at risk of developing CRC compared to healthy subjects, indicating a potential association between these three major SCFA molecules and CRC development. This finding was further confirmed in the CRC incidence category analysis where the faecal levels of SCFAs in CRC patients were significantly lower compared to those in healthy subjects.

Our findings in CRC risk and incidence were consistent with the observations reported in other meta-analyses, which focused on the association between dietary fibre intake and the risk of colorectal adenoma (Nucci *et al*. 2021; Oh *et al*. 2019), and carcinoma (Gianfredi *et al*. 2018). These systematic reviews suggested a protective effect of dietary fibre intake against CRA and CRC (Gianfredi *et al*. 2018; Nucci *et al*. 2021; Oh *et al*. 2019). Since SCFAs are produced by gut-microbiota via the fermentation of dietary fibres (Alexander *et al*. 2019; Liu *et al*. 2021; Parada Venegas *et al*. 2019; van der Beek *et al*. 2017), our meta-analysis of SCFA concentrations in CRC further confirms earlier observations and underlines the importance of dietary fibres/SCFAs in the risk and progression of CRC.

Another meta-analysis, which assessed the effect of non-digestible carbohydrate [resistance starch (RS)] or inulin supplementation on the risk of colorectal neoplasia, did not find significant increase in fecal total SCFA or butyric acid concentration and excretion before and after the intervention (Rao *et al*. 2021). Many studies which investigated the effect of RS on healthy subjects or individuals with sporadic CRC or adenoma had a period of ≤ 4-week of intervention. A few studies reported 7- and 8-week intervention on adenoma or healthy individuals and the remaining studies were conducted on individuals with inherited CRC syndromes after > 2-year intervention (Rao *et al*. 2021). The duration of intervention was longest (> 2 years) for studies involving hereditary CRC cases with reported germ-line mutations, which may have outweighed the effect of RS supplementation. While interventions involving sporadic cases or healthy subjects had much shorter periods of RS intervention (< 8 weeks) (Rao *et al*. 2021). In our meta-analysis, we also did not observe a significant difference in total fecal SCFAs in the CRC risk category. This could be due to other SCFA molecules such as valeric, iso-butyric, and iso-valeric acid being included in total SCFA measurements; the latter two are the branched SCFAs mainly produced via fermentation of branched amino acids in the colon and not from non-digestible carbohydrates (Gill *et al*. 2018; Rios-Covian *et al*. 2020).

Another systematic review on the food-microorganism-SCFA axis, without any meta-analysis, concluded that most evidence demonstrated higher SCFA levels in individuals at risk of CRC compared to healthy individuals (Shuwen *et al*. 2019), which contrasts with findings in our systematic review which showed lower fecal SCFA concentration in at-risk individuals (**Figure 2**). In comparison to our systematic review, their search strategy restricted their analysis to only 8 of the final 22 studies that we analysed (Bridges *et al*. 2018; Hester *et al*. 2015; S. J. D. O’Keefe *et al*. 2009; Ohigashi *et al*. 2013; Ou *et al*. 2013; Ou *et al*. 2012; Weaver *et al*. 1988; Weir *et al*. 2013). Therefore, their conclusion was based on a smaller subset of the primary studies available and was also not supported by a meta-analysis.

Both the quantitative and qualitative analyses of CRC risk identified comparable findings of significantly lower concentration of acetic and butyric acid in the high-versus low-risk CRC group. For the CRC incidence category, the quantitative meta-analysis of butyric acid was consistent with observations identified in most of the articles from the qualitative analysis, supporting the evidence of lower concentration of three SCFAs in CRC cases compared to healthy controls. The meta-analysis of propionic acid was not significantly different between cases and controls. Similarly, most of the studies (4 of 6) reported no significant difference in fecal propionic acid concentration between CRC and healthy control in the qualitative analysis. The meta-analysis on IBS revealed a significantly higher concentration of fecal propionic acid in these patients in comparison to healthy controls (Sun *et al*. 2019). Therefore, further studies comparing SCFA profiles among multiple gut diseases could shed more light on the importance of these molecules in the development of varied medical conditions.

To our knowledge, this systematic review is the first to provide a comprehensive search and data collection on observational studies linking SCFA molecules with the CRC risk and incidence. A limitation of our analysis is the heterogeneity of the studies evaluated in this systematic review, which is very difficult to control for. One such factor was the age group assessed for CRC incidence and risk. The mean age of the group in the studies was greater than 50 years and fecal SCFA concentration was not measured in younger populations to provide a comparison with low-risk, young age individuals. Although CRC is most often diagnosed in individuals > 50 years, the incidence for early-onset CRC (EOCRC) in adults aged 20-49 years has increased over the past decade in the USA, Australia, and Europe (Akimoto *et al*. 2021; Burnett-Hartman *et al*. 2021; Saad El Din *et al*. 2020; Vuik *et al*. 2019). It would be of interest in the future to study different age group populations for CRC risk and incidence. Family history (Kastrinos, Samadder and Burt 2020), diet and lifestyle (Baena and Salinas 2015) are known factors contributing to CRC incidence. Only a few studies assessed in this systematic review provided information on the dietary difference between groups (Bridges *et al*. 2018; H. M. Chen *et al*. 2013; Katsidzira *et al*. 2019; S. J. D. O’Keefe *et al*. 2009; Ocvirk *et al*. 2020).

Another limitation could arise from the different methodologies used to measure fecal SCFA across the studies (**Table 1**). This systematic review did not include non-English records. To our knowledge, no longitudinal studies have reported fecal SCFA measurements at different time points during CRC progression, nonetheless, the 22 studies assessed in this systematic review and meta-analysis provide a comparison between CRC risk/incidence and respective controls from various countries and ethnic groups.

In addition to the SCFAs assessed in this systematic review, other metabolites such as bile acids were also measured in six of the selected studies (C. Y. Chen *et al*. 2021; Katsidzira *et al*. 2019; Ocvirk *et al*. 2020; Ou *et al*. 2012; Torii, Kanemitsu and Hagiwara 2019; Weir *et al*. 2013). Among the bile acids investigated, a significantly higher fecal concentration of deoxycholic acid in the CRC high-versus low-risk group was reported in three studies (Katsidzira *et al*. 2019; Ocvirk *et al*. 2020; Ou *et al*. 2012). Dietary fibre and fat promote the production of SCFA and bile acid molecules in the gut, respectively, and the latter is associated with gastrointestinal carcinogenesis (Jia, Xie and Jia 2018; T. T. Nguyen *et al*. 2018; Ocvirk *et al*. 2019). Measurement of fecal SCFAs and other gut metabolites (such as bile acids) in longitudinal studies comparing individuals with colorectal adenoma/risk and healthy subjects could strengthen their association with CRC progression.

## Conclusion

Gut microbiota dysbiosis and changes in their metabolites have been the focus of epidemiological studies aimed at uncovering associations with colonic inflammation and carcinogenesis. In line with the protective role of fecal SCFAs against the development of gut diseases (Liu *et al*. 2021; Parada Venegas *et al*. 2019), and the protective effect of dietary fibres against CRC risk and/or incidence (Gianfredi *et al*. 2018; Nucci *et al*. 2021; Oh *et al*. 2019), we determined that the combined fecal concentration of the three major SCFA molecules was significantly lower not only in CRC patients compared to healthy controls, but also in high-risk CRC individuals.

This study supports further exploration into fecal concentration of SCFAs: acetic, propionic, and butyric acids, as biomarkers for CRC risk. Among the current CRC screening methods, colonoscopy is the gold standard (Davidson *et al*. 2021) however, being invasive it presents some procedural risk (Ferrari *et al*. 2021). The guaiac fecal occult blood test (gFOBT) and fecal immunochemical test (FIT) are other, in practice, non-invasive stool-based methods for CRC screening, which however require improvement, in particular for detection of CRA or early-stage colonic carcinogenesis (Ferrari *et al*. 2021; Imperiale *et al*. 2019; Jodal *et al*. 2019). Fecal SCFA could be considered as a potential non-invasive biomarker to be measured in combination with or as an alternative to the commonly used non-invasive and current CRC screening methods (Anghel *et al*. 2021; Ferrari *et al*. 2021), to improve specificity and sensitivity of current screening, as well as for potential early detection of CRA.

## Registration

The study is registered in PROSPERO database (registration code: CRD42021256123).

## Data Availability

All data produced in the present work are contained in the manuscript

## Acknowledgements

Authors acknowledge Ho Pham for his assistance and the infrastructure support through the School of Medicine, Western Sydney University. Authors also acknowledge Ameneh Najdi for checking the data entry in the analyses.

## Author contributions

Conceptualization: AAH; Original search and analysis: EA; Validation: WKMW; Writing-first draft: EA; Writing-review and editing: WKMW, MVJ, KS, AAH; Writing- finalisation: KS and AAH. KS and AAH are the guarantors of this work and, as such, had full access to all the data in the study and take responsibility for the integrity of the data and the accuracy of the data analysis.

## Supplementary Materials

### Supplementary Methods

The details of the search strategy conducted on May 21, 2021.

<u>1) Medline and Embase combined search, followed by deduplication, using Ovid search interface: Search link:</u>

https://ezproxy.uws.edu.au/login?url=http://ovidsp.ovid.com/ovidweb.cgi?T=JS&xsNEWS=N&PAGE=main&SHAREDSEARCHID=6oHhfLqbyqUT2KwL2FpUgcWyfohUdPU40kumYqdCTlI8X2Ibo6ZjVT3zsbLZCbIw7

Search history details:

Embase <1974 to 2021 May 20>

Ovid MEDLINE(R) ALL

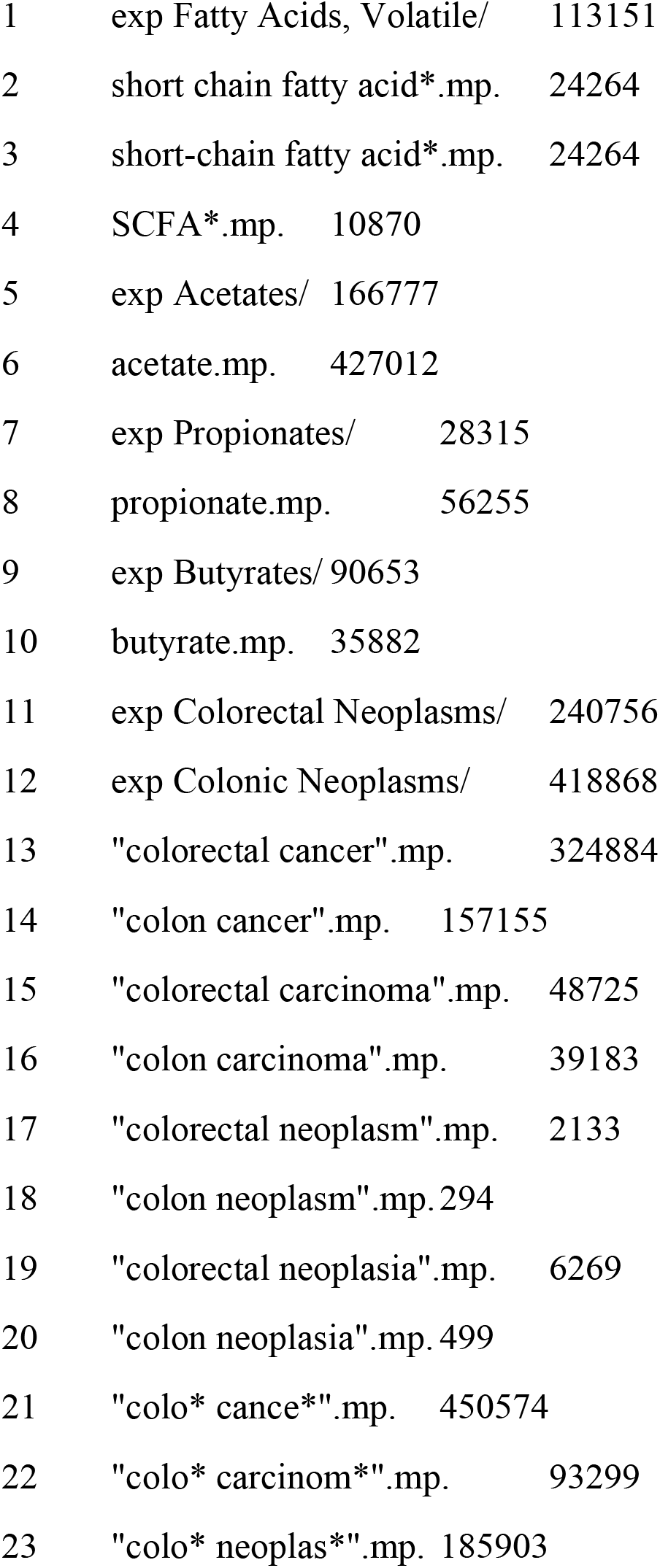
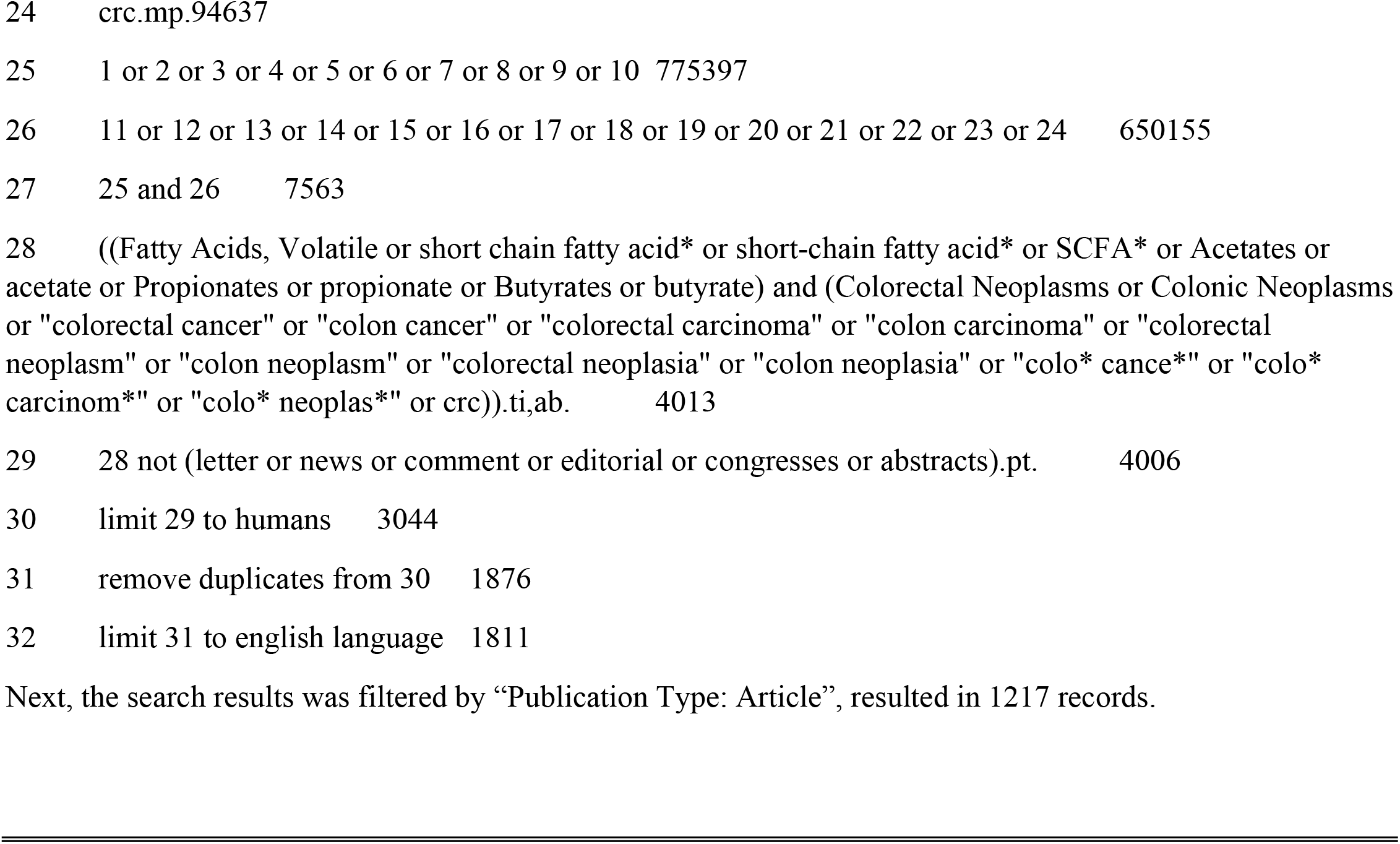

<u>2) Web of science:</u>

ab=((“scfa*” OR “short chain fatty acid*” OR “short-chain fatty acid*” OR acetate OR propionate OR butyrate) AND (“colo* cance*” OR “colo* carcinom*” OR “colo* neoplas*” OR crc) AND (human* OR patient* OR subject* OR individual* OR participant* OR case* OR control* OR character* person* OR people) NOT (rats OR mouse OR mice OR murine))

Next, the search results were refined by “Document Types: Articles”, and “Languages: English”, resulted in 783 records.

**Supplementary Table 1.** Quality assessment of the selected case-control studies (n = 16) using Newcastle-Ottawa Scale (NOS). According to NOS guideline, one star can be given to each sector in Selection or Exposure category, and two stars for Comparability category, one per one matched factor. ^¶^Refer to the text for the definition of CRC incidence or risk category. ^§^Based on the definition of control in NOS guideline, the star was given only to studies that “explicitly stated that controls have no history of this outcome”.

|  |  |  | Selection |  |  |  | Comparability | Exposure |  |  |  |
| --- | --- | --- | --- | --- | --- | --- | --- | --- | --- | --- | --- |
|  | Study | Analysis Category<sup>¶</sup> | Adequate definition of cases | Representativeness of cases | Selection of controls | Definition of controls<sup>§</sup> | Control for important factor or additional factor | Ascertainment of exposure | Same method of ascertainment for cases and controls | Non-response rate | Total scores |
| Qualitative data<br>(values of SCFA levels not | Sze et al. 2019 | Incidence and risk | ★ | ★ |  |  |  | ★ | ★ | ★ | 5 |
|  | Lin et al. 2019 | Incidence | ★ |  | ★ |  | ★★ | ★ | ★ | ★ | 7 |
|  | Lin et al. 2016 | Incidence | ★ | ★ | ★ |  |  | ★ | ★ | ★ | 6 |
|  | Weir et al. 2013 | Incidence | ★ |  |  |  |  | ★ | ★ | ★ | 4 |
|  | Ohigashi et al. 2013 | Incidence and risk | ★ | ★ | ★ |  |  | ★ | ★ | ★ | 6 |
|  | Monleon et al. 2009 | Incidence | ★ | ★ | ★ |  |  | ★ | ★ | ★ | 6 |
|  | Weaver et al. 1988 | Incidence | ★ | ★ | ★ | ★ |  | ★ | ★ | ★ | 7 |
| Quantitative data<br>(reported SCFA values) | Chen et al. 2021 | Risk |  | ★ | ★ |  |  | ★ | ★ | ★ | 5 |
|  | Torii et al. 2019 | Incidence | ★ |  |  |  |  | ★ | ★ | ★ | 4 |
|  | Niccolai et al. 2019 | Incidence and risk | ★ | ★ | ★ |  |  | ★ | ★ | ★ | 6 |
|  | Yusuf et al. 2018 | Incidence | ★ | ★ | ★ |  |  | ★ | ★ | ★ | 6 |
|  | Song et al. 2018 | Incidence | ★ | ★ | ★ | ★ | ★ | ★ | ★ | ★ | 8 |
|  | Bridges et al. 2018 | Risk | ★ | ★ | ★ | ★ |  | ★ | ★ | ★ | 7 |
|  | Chen et al. 2013 | Risk | ★ | ★ | ★ |  |  | ★ | ★ | ★ | 6 |
|  | Boutron-Ruault et al. 2005 | Risk | ★ | ★ | ★ | ★ | ★★ | ★ | ★ | ★ | 9 |
|  | Kashtan et al. 1992 | Risk | ★ | ★ | ★ |  |  | ★ | ★ | ★ | 6 |

**Supplementary Table 2.** Quality assessment of the included cross-sectional studies (n = 6) using Joanna Briggs Institute (JBI) Critical Appraisal tool. The studies are shown with alternate shading. ^¶^Matched for sex. ^§^Matched for age and sex. ^†^Matched for age, as recruited subjects aged 50-60 years old.

| Ocvirk et al. 2020 | Response options |  |  |  |
| --- | --- | --- | --- | --- |
| 1. Were the criteria for inclusion in the sample clearly defined? | <u>Yes</u> | No | Unclear | Not applicable |
| 2. Were the study subjects and the setting described in detail? | <u>Yes</u> | No | Unclear | Not applicable |
| 3. Was the exposure measured in a valid and reliable way? | <u>Yes</u> | No | Unclear | Not applicable |
| 4. Were objective, standard criteria used for measurement of the condition? | <u>Yes</u> | No | Unclear | Not applicable |
| 5. Were confounding factors identified? | <u>Yes</u> | No | Unclear | Not applicable |
| 6. Were strategies to deal with confounding factors stated? | Yes | <u>No</u> | Unclear | Not applicable |
| 7. Were the outcomes measured in a valid and reliable way? | <u>Yes</u> | No | Unclear | Not applicable |
| 8. Was appropriate statistical analysis used? | <u>Yes</u> | No | Unclear | Not applicable |
| Overall appraisal: Include ☒ Exclude ☐ Seek further info ☐ |  |  |  |  |
| Katsidzira et al. 2019 | Response options |  |  |  |
| 1. Were the criteria for inclusion in the sample clearly defined? | <u>Yes</u> | No | Unclear | Not applicable |
| 2. Were the study subjects and the setting described in detail? | <u>Yes</u> | No | Unclear | Not applicable |
| 3. Was the exposure measured in a valid and reliable way? | <u>Yes</u> | No | Unclear | Not applicable |
| 4. Were objective, standard criteria used for measurement of the condition? | <u>Yes</u> | No | Unclear | Not applicable |
| 5. Were confounding factors identified? | <u>Yes</u> | No | Unclear | Not applicable |
| 6. Were strategies to deal with confounding factors stated? | <u>Yes</u><sup>¶</sup> | No | Unclear | Not applicable |
| 7. Were the outcomes measured in a valid and reliable way? | <u>Yes</u> | No | Unclear | Not applicable |
| 8. Was appropriate statistical analysis used? | <u>Yes</u> | No | Unclear | Not applicable |
| Overall appraisal: Include ☒ Exclude ☐ Seek further info ☐ |  |  |  |  |
| Hester et al. 2015 | Response options |  |  |  |
| 1. Were the criteria for inclusion in the sample clearly defined? | <u>Yes</u> | No | Unclear | Not applicable |
| 2. Were the study subjects and the setting described in detail? | Yes | <u>No</u> | Unclear | Not applicable |
| 3. Was the exposure measured in a valid and reliable way? | <u>Yes</u> | No | Unclear | Not applicable |
| 4. Were objective, standard criteria used for measurement of the condition? | Yes | <u>No</u> | Unclear | Not applicable |
| 5. Were confounding factors identified? | <u>Yes</u> | No | Unclear | Not applicable |
| 6. Were strategies to deal with confounding factors stated? | Yes | <u>No</u> | Unclear | Not applicable |
| 7. Were the outcomes measured in a valid and reliable way? | Yes | No | <u>Unclear</u> | Not applicable |
| 8. Was appropriate statistical analysis used? | <u>Yes</u> | No | Unclear | Not applicable |
| Overall appraisal: Include ☒ Exclude ☐ Seek further info ☐ |  |  |  |  |

| Ou et al. 2013 | Response options |  |  |  |
| --- | --- | --- | --- | --- |
| 1. Were the criteria for inclusion in the sample clearly defined? | <u>Yes</u> | No | Unclear | Not applicable |
| 2. Were the study subjects and the setting described in detail? | <u>Yes</u> | No | Unclear | Not applicable |
| 3. Was the exposure measured in a valid and reliable way? | <u>Yes</u> | No | Unclear | Not applicable |
| 4. Were objective, standard criteria used for measurement of the condition? | <u>Yes</u> | No | Unclear | Not applicable |
| 5. Were confounding factors identified? | <u>Yes</u> | No | Unclear | Not applicable |
| 6. Were strategies to deal with confounding factors stated? | <u>Yes</u><sup>s</sup> | No | Unclear | Not applicable |
| 7. Were the outcomes measured in a valid and reliable way? | <u>Yes</u> | No | Unclear | Not applicable |
| 8. Was appropriate statistical analysis used? | <u>Yes</u> | No | Unclear | Not applicable |
| Overall appraisal: Include ☒ Exclude ☐ Seek further info ☐ |  |  |  |  |
| Ou et al. 2012 | Response options |  |  |  |
| 1. Were the criteria for inclusion in the sample clearly defined? | <u>Yes</u> | No | Unclear | Not applicable |
| 2. Were the study subjects and the setting described in detail?<br>(According to this reference of the paper: O'Keefe et al. 2007, PMID: 17182822.) | <u>Yes</u> | No | Unclear | Not applicable |
| 3. Was the exposure measured in a valid and reliable way? | <u>Yes</u> | No | Unclear | Not applicable |
| 4. Were objective, standard criteria used for measurement of the condition? | <u>Yes</u> | No | Unclear | Not applicable |
| 5. Were confounding factors identified? | <u>Yes</u><sup>†</sup> | No | Unclear | Not applicable |
| 6. Were strategies to deal with confounding factors stated? | Yes | <u>No</u> | Unclear | Not applicable |
| 7. Were the outcomes measured in a valid and reliable way? | <u>Yes</u> | No | Unclear | Not applicable |
| 8. Was appropriate statistical analysis used? | <u>Yes</u> | No | Unclear | Not applicable |
| Overall appraisal: Include ☒ Exclude ☐ Seek further info ☐ |  |  |  |  |
| O'Keefe et al. 2009 | Response options |  |  |  |
| 1. Were the criteria for inclusion in the sample clearly defined? | <u>Yes</u> | No | Unclear | Not applicable |
| 2. Were the study subjects and the setting described in detail?<br>(Only the time period of living in either area wasn't mentioned.) | <u>Yes</u> | No | Unclear | Not applicable |
| 3. Was the exposure measured in a valid and reliable way?<br>(According to this reference of the paper: O'Keefe et al. 2007, PMID: 17182822.) | <u>Yes</u> | No | Unclear | Not applicable |
| 4. Were objective, standard criteria used for measurement of the condition? | <u>Yes</u> | No | Unclear | Not applicable |
| 5. Were confounding factors identified? | <u>Yes</u><sup>†</sup> | No | Unclear | Not applicable |
| 6. Were strategies to deal with confounding factors stated? | Yes | <u>No</u> | Unclear | Not applicable |
| 7. Were the outcomes measured in a valid and reliable way? | <u>Yes</u> | No | Unclear | Not applicable |
| 8. Was appropriate statistical analysis used? | <u>Yes</u> | No | Unclear | Not applicable |
| Overall appraisal: Include ☒ Exclude ☐ Seek further info ☐ |  |  |  |  |

**Supplementary Table 3.** Table showing which data is used for which analysis. Cross-sectional studies are highlighted in grey, and case-control studies are not highlighted. Total number of studies in each category is stated at the bottom of table. Note: for quantitative analyses in risk category, the meta-analyses in the **Figure 2** included 3, 3, 5, and 3 studies for measuring C2, C3, C4, and total SCFA, respectively. C2: acetic acid, C3: propionic acid, C4: butyric acid.

|  |  | CRC risk |  |  |  | CRC incidence |  |  |  |
| --- | --- | --- | --- | --- | --- | --- | --- | --- | --- |
|  |  | C2 | C3 | C4 | Total SCFA | C2 | C3 | C4 | Total SCFA |
| Qualitative data (values of SCFA levels not reported) | Sze et al. 2019 | * | * | * |  | * | * | * |  |
|  | Lin et al. 2019 |  |  |  |  | * | * | * |  |
|  | Lin et al. 2016 |  |  |  |  | * | * | * |  |
|  | Weir et al. 2013 |  |  |  |  | * | * | * |  |
|  | Ohigashi et al. 2013 | * |  |  |  | * |  |  |  |
|  | Monleon et al. 2009 |  |  |  |  | * | * | * |  |
|  | Weaver et al. 1988 |  |  |  |  | * | * | * | * |
|  | Ocvirk et al. 2020 | * | * |  | * |  |  |  |  |
|  | Katsidzira et al. 2019 | * | * | * |  |  |  |  |  |
|  | Hester et al. 2015 | * | * | * | * |  |  |  |  |
|  | Ou et al. 2013 | * | * | * |  |  |  |  |  |
|  | Ou et al. 2012 | * | * | * |  |  |  |  |  |
|  | O'Keefe et al. 2009 | * | * | * | * |  |  |  |  |
| Quantitative data (reported SCFA values) | Chen et al. 2021 | * | * | * |  |  |  |  |  |
|  | Torii et al. 2019 |  |  |  |  | * | * | * |  |
|  | Niccolai et al. 2019 | * | * | * | * | * | * | * | * |
|  | Yusuf et al. 2018 |  |  |  |  | * | * | * |  |
|  | Song et al. 2018 |  |  |  |  | * | * | * |  |
|  | Bridges et al. 2018 | * | * | * | * |  |  |  |  |
|  | Chen et al. 2013 | * | * | * |  |  |  |  |  |
|  | Boutron-Ruault et al. 2005 | * | * | * | * |  |  |  |  |
|  | Kashtan et al. 1992 |  |  | * | * |  |  |  |  |
|  | Ocvirk et al. 2020 |  |  | * |  |  |  |  |  |
|  | Total number of studies, providing **qualitative** data | 8 | 7 | 6 | 3 | 7 | 6 | 6 | 1 |
|  | Total number of studies, providing **quantitative** data | 5 | 5 | 7 | 4 | 4 | 4 | 4 | 1 |
|  |  | C2 | C3 | C4 | Total SCFA | C2 | C3 | C4 | Total SCFA |
|  |  | CRC risk |  |  |  | CRC incidence |  |  |  |

**Supplementary Table 4.** The summary of other systematic reviews related to fibre intake, SCFA and risk of colorectal cancer or adenoma.

| Study | PMID | Title | Aim | Primary source | Meta-analysis | Outcome |
| --- | --- | --- | --- | --- | --- | --- |
| Rao et al. 2021 | 32202158 | Non-Digestible Carbohydrate and the Risk of Colorectal Neoplasia: A Systematic Review | To check whether RS and inulin should be offered to cancer/precancerous patients or healthy subjects to decrease their risk of CRC. | Interventional studies on resistant starch (RS) or inulin supplementation | Total SCFA and butyrate concentration and excretion | Total SCFAs and butyrate concentrations and excretions in feces did not increase significantly after RS/inulin supplementation. |
| Nucci et al. 2021 | 33920845 | Association between Dietary Fibre Intake and Colorectal Adenoma: A Systematic Review and Meta-Analysis | Assessing the association between dietary fibre intake and the risk of colorectal adenoma in adults. | Interview and questionnaire-based case-control, cohort, and cross-sectional studies | Effect size is measured as odds ratio | There could be a protective effect of dietary fibre intake against colorectal adenoma. |
| Shuwen et al. 2019 | 31401674 | Protective effect of the "food-microorganism-SCFAs" axis on colorectal cancer: from basic research to practical application | To elucidate the "food-microorganism-SCFAs" axis and to provide guidance for prevention and intervention in CRC. | Not specific. Human, animal, and cell-based studies | - | The concentrations of SCFAs in CRC patients and individuals with a high risk of CRC were higher than those in healthy individuals. |
| Oh et al. 2019 | 31495339 | Different dietary fibre sources and risks of colorectal cancer and adenoma: a dose-response meta-analysis of prospective studies | To summarise the relationships of different fibre sources with colorectal cancer and adenoma risks. | Interview and questionnaire-based prospective cohort studies | Effect size is measured as relative risk | The evidence for colorectal cancer prevention is strongest for fibre from cereals/grains. Each 10 g/d increase in dietary intake of vegetable or fruit fibre was statistically significantly associated with a reduced risk of incident colorectal adenoma. |
| Gianfredi et al. 2018 | 29516760 | Is dietary fibre truly protective against colon cancer? A systematic review and meta-analysis | To evaluate the association between dietary fibre intake and the risk of colon cancer. | Interview and questionnaire-based case-control, cohort, and cross-sectional studies | Effect size is measured as odds ratio | Results suggest a protective role of dietary fibre intake on colon cancer risk. |

**Supplementary Figure 1.**
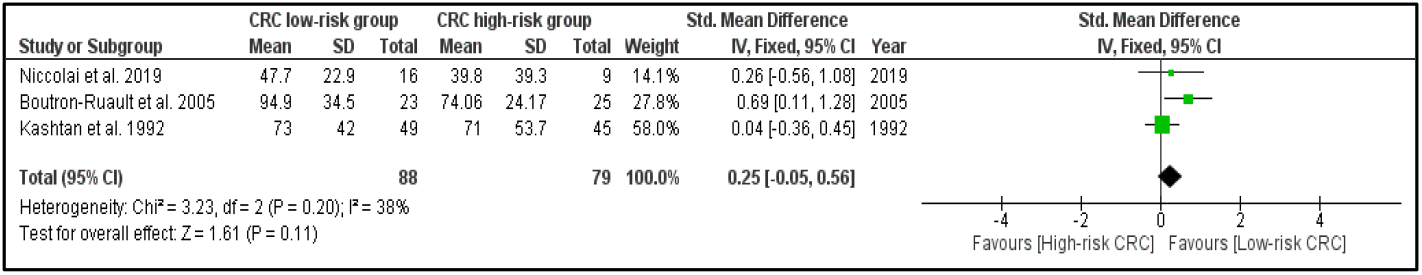
Forest plot representing the meta-analyses of the fecal total SCFA concentration in the CRC risk category using fixed-effect model. Note that total SCFA indicates the collection of all the SCFA molecules - not only acetic, propionic, and butyric acid.

**Supplementary Figure 2.**
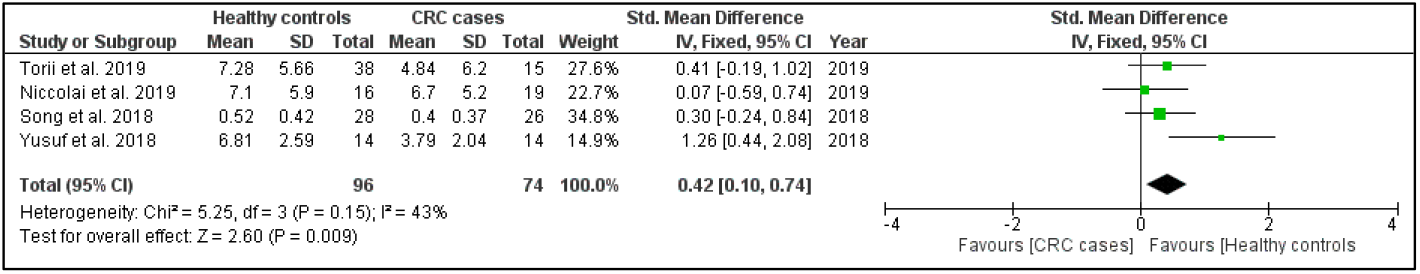
Forest plot representing the meta-analyses of the fecal butyric acid concentration in the CRC incidence category using fixed-effect model.

**Supplementary Figure 3.**
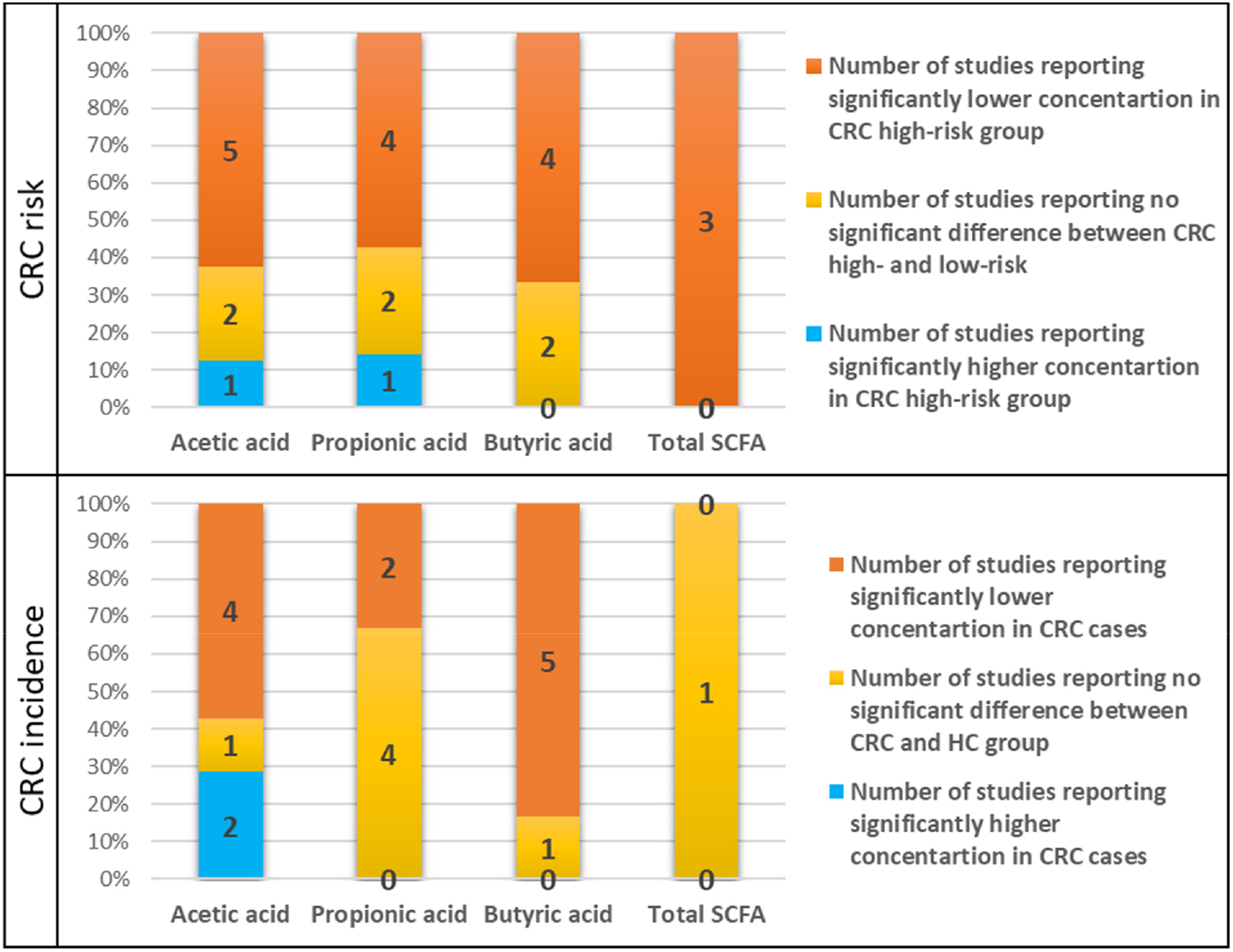
Graphical representation of fecal SCFA concentration. Stacked bar charts summarizing the results of the qualitative data in CRC risk (8, 7, 6, and 3 studies have measured fecal concentration of acetic, propionic, and butyric acid, and total SCFA, respectively) and incidence categories (7, 6, 6, and 1 study have measured fecal concentration of acetic, propionic, and butyric acid, and total SCFA, respectively). HC: healthy controls.

